# Combination of QTL and GWAS to uncover the role of phosphodiesterases in ischemic heart disease

**DOI:** 10.1101/2023.09.24.23296052

**Authors:** Jun Xiao, Ziting Gao, Hongye Wei, Yajing Wei, Ziyi Qiu, Wuqing Huang

**Author notes:** **Corresponding author:** Wuqing Huang, Fujian Medical University, No 1, Xue Yuan Road, University Town, 350108 Fuzhou City, Fujian Province, China.

## Abstract

**Background:** Phosphodiesterases (PDEs) are regarded as important therapeutic targets for multiple diseases, and the cardiovascular benefits of several PDE inhibitors have received extensive interests.

**Objectives:** To explore the relationship between genetically-predicted PDEs and ischemia heart disease via drug target Mendelian Randomization (MR) approach.

**Methods:** The expression of genes encoding PDEs was used to proxy the level of PDEs, and available quantitative trait loci of gene expression and DNA methylation (eQTLs and mQTLs) for each target gene were identified as the genetic instruments. Coronary heart disease (CHD) and myocardial infarction (MI) were the outcomes. Summary-data-based MR method was used to generate the estimates and two-step MR analysis was applied to investigate the mediation of metabolic traits.

**Results:** MR analyses identified two types of PDEs, PDE5 and PDE8, genetically-predicted expression in blood of the encoded genes was significantly associated with the risk of CHD (OR_PDE5A_=1.22,95% CI=1.06-1.40; OR_PDE8A_=1.26,95% CI=1.07-1.49) and MI (OR_PDE5A_=1.27,95% CI=1.09-1.48; OR_PDE8A_=1.24,95% CI=1.04-1.48). Especially, the highest expression of PDE5A was observed in artery aorta, which was also positively related to CHD (OR=1.17,95% CI=1.05-1.32) and MI (OR=1.15,95% CI=1.02-1.30). Besides, the methylation level of 12 CpG sites showed a relation with CHD or MI via affecting PDE5A expression. The observed association between PDE5A expression and outcomes were partly mediated by blood pressure and LDL cholesterol, and the association with MI were mostly mediated by CHD (Proportion-mediated: 78.84%).

**Conclusions:** This study provided genetic evidence about the protective role of PDE5 inhibition against ischemic heart disease, especially in preventing patients with CHD from developing MI.

## Introduction

Phosphodiesterases (PDEs), a family of enzymes from PDE1 to PDE11, are well-known to play a critical role in the regulation of intracellular signaling via hydrolyzing cyclic nucleotides cyclic adenosine monophosphate (cAMP) and/or cyclic guanosine monophosphate (cGMP)(1). The cAMP and cGMP are the second messengers linking to a series of physiological processes, thus PDEs are regarded as important therapeutic targets for multiple diseases (2). Although the PDE families are structurally related, the molecular diversity makes them functionally distinct (2). Some PDE inhibitors have been widely used in clinical practice, and emerging evidence indicates that PDE inhibitors have the potential to be developed or repurposed for a wide range of clinical applications (2,3).

The cardiovascular benefits of PDE inhibitors have received extensive interests in recent decades, for example, accumulating epidemiological studies suggested that PDE5 inhibitors could enhance contractile function for patients with heart failure, improve the prognosis in patients with coronary heart disease, reduce ventricular arrhythmias, and so on (4–16). Coronary heart disease (CHD) is the most common type of cardiovascular diseases, and myocardial infarction (MI) is a major cause of death worldwide. Since 1960s, cAMP and cGMP were found to inhibit platelet aggregation and a group of PDE inhibitors were considered as anti-platelet agents, for instance, dipyridamole (PDE3 inhibitor) has been approved for secondary prevention of ischemic stroke (17). PDE inhibitors were also reported to ameliorate some established risk factors of coronary heart disease, such as metabolic syndrome, a cluster of several metabolic disorders (18). Although these evidence have suggested the cardiovascular benefits of PDE inhibitors, there is a paucity of studies comprehensively exploring the causal effect of PDE inhibitors on CHD or MI.

Randomized clinical trial (RCT) is recognized as the gold standard to confirm the causal effect, however, RCT is not always feasible due to the difficulty of implementation. Mendelian randomization (MR) is thus developed for causal inference by using genetic variants as instruments. Previous studies have applied drug-target MR method to explore the potential drug targets by using quantitative trait loci (QTLs) as instruments, which are genetic variants associated with quantitative traits, such as gene expression (eQTL), protein (pQTL), methylation (mQTL), et al(19,20).

Therefore, in this study, we aimed to investigate the role of PDEs on CHD and MI by using available eQTLs and mQTLs of genes encoding PDEs as instruments via MR approach and explore how the identified PDEs affect the outcome through metabolic traits.

## Methods

### Study design

The design of the study was summarized in **Figure 1**. In the primary analysis, we comprehensively investigated the causal effect of the expression of genes encoding PDEs on the risk of CHD or MI by using eQTLs from blood as instruments. For significant MR associations, we further conducted a series of secondary analyses. Firstly, the observed significant associations were examined by using eQTLs in artery or heart tissues as instruments. Secondly, it is known that DNA methylation plays an important role in regulating gene expression, thus we further investigated the relationship between methylation level of related genes encoding significant PDEs and the outcomes by using mQTLs in blood as instruments. Thirdly, two-step MR analysis was used to explore if metabolic traits perform a mediating role between the identified PDEs and the outcomes, including body mass index (BMI), blood pressure, blood lipids and blood glucose.

**Figure 1.**
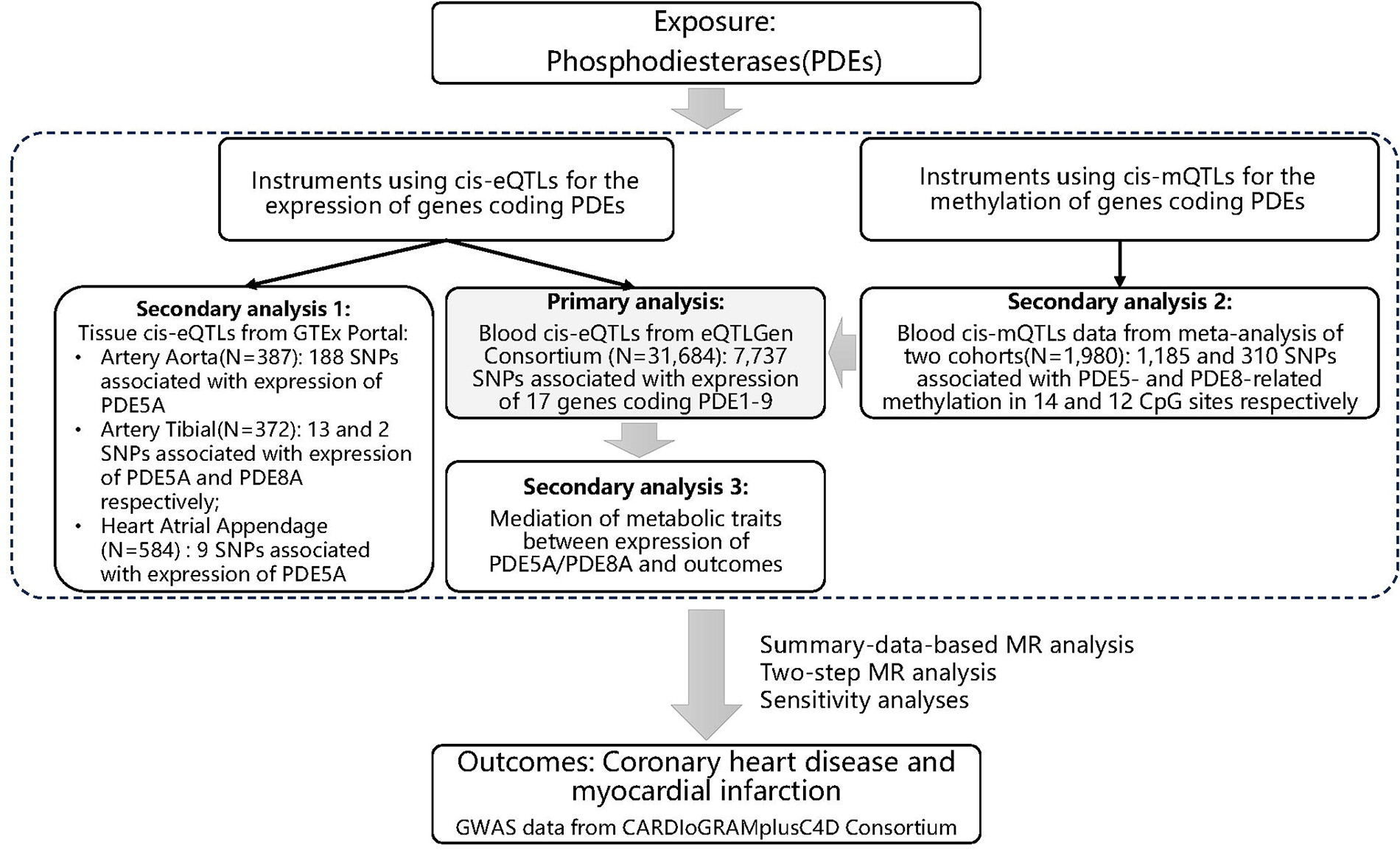
Flowchart of the study.

Summary-level data from publicly available genome-wide association studies (GWASs), expression and methylation quantitative trait loci (eQTLs and mQTLs) studies, were used in this study, all of which have been approved by the relevant institutional review boards with informed consents from participants. Details of data resources are presented in **Supplementary Table 1**.

### Selection of genetic instruments

We used the expression of the encoded gene to proxy the level of each PDE, and eQTLs for related genes were used as instruments of gene expression. Given that data of eQTLs in blood are most comprehensive and top eQTLs are highly correlated between blood and other tissues, eQTLs in blood were used to generate genetic instruments to identified significant PDEs in the primary analysis. For significant associations, we provided additional evidence by using eQTLs in disease-related organs (ie.,artery and heart tissues) as instruments. Summary-level data of eQTLs in blood and tissues were obtained from eQTLGen Consortium (N=31,684) and GTEx Consortium V8 (Artery Aorta: N=387; Artery Tibial: N=372; Heart Atrial Appendage: N=584), respectively (21,22). And common SNPs significantly associated with the expression and the methylation in blood for related genes were identified and used to generate instruments of the level of methylation, summary data of mQTLs were selected from a meta-analysis of two cohorts (N=1,980) (23). To reduce weak instrument bias, we only included cis-QTLs (i.e., QTLs within 1LJMb windows from the region of the encoded gene). Hence, common cis-eQTLs or cis-mQTLs (minor allele frequency (MAF) >1%) significantly associated with the expression or methylation of PDEs were identified, in which p<5.0LJ×LJ10^-8^ was set as the significant threshold to select cis-eQTLs or cis-mQTLs in blood and the threshold of p<1.0LJ×LJ10^-5^ was used to select cis-eQTLs in tissues due to the limited number of available cis-eQTLs in tissues and the relatively small sample size of original studies. The most significant cis-eQTL or cis-mQTL for each gene was finally selected as the genetic instrument. The coefficient obtained from eQTL or mQTL study is equivalent to 1-SD change of the level of gene expression or DNA methylation for each additional effect allele. The details of QTLs data are presented in **Supplementary Table 1.**

### Outcome sources

The outcome is ischemic heart disease, including CHD and MI. The GWAS summary-level data for CHD (cases: N=60,801; controls: N=123,504) and MI (cases: N=43,676; controls: N=128,199) were obtained from the CARDIoGRAMplusC4D Consortium with meta-analyses of 48 GWASs, in which the study population was predominantly individuals with European ancestry (24). Metabolic traits were considered as the potential mediators: summary data of GWAS for systolic and diastolic blood pressure was obtained from the International Consortium of Blood Pressure; data for low-density lipoprotein cholesterol (LDL-C), high-density lipoprotein cholesterol (HDL-C) and triglycerides (TG) obtained from the Global Lipids Genetics Consortium; data for body mass index obtained from the GIANT Consortium; data for fasting blood glucose obtained from the MAGIC Consortium. The details of GWASs data are presented in Supplementary Table 1.

### Statistical analyses

Effect estimates were generated by using summary-data-based MR (SMR) method, which was recently developed to evaluate the association of quantitative trait and outcome combining summary-level data from QTL and GWAS studies (25). F-statistic was calculated to assess the strength of genetic instruments, an F-statistic of >10 indicates a small role of weak instrument bias in the results (26). The heterogeneity in dependent instruments (HEIDI) test was applied to assess if linkage disequilibrium played a role in the observed association of gene expression or methylation with outcome of interest, in which p value <0.01 indicates that association may be due to linkage. One SNP probably regulates the expression of more than one genes, resulting in the presence of horizontal pleiotropy. To evaluate the potential impact of horizontal pleiotropy, genes near the instrumental variant (within a 1Mb window) were identified if the expression was significantly associated with the variant. Then SMR analysis was conducted to examine if the instrumental variant affected the risk of outcomes via regulating the expression of nearby genes.

Inverse-variance weighted mendelian randomization (IVW-MR) method was applied to obtain the estimates of the causal associations between metabolic traits and the outcome. To evaluate if the significant association between PDE and outcome was mediated by metabolic traits, we conducted a two-step MR study between PDE, candidate mediators, and incident outcome. Two-step MR, also known as network MR, is similar to the approach calculating the product of coefficients (27). We first calculated two MR coefficients: the association between PDE and the mediator (beta1) and the association between the mediator and the outcome (beta2), then multiplied two coefficients to estimate the indirect effect (beta1*beta2). The total effect was estimated by MR coefficient of PDE on the outcome (beta0) and the direct effect was calculated by the difference between total effect and indirect effect (beta0-beta1*beta2). Proportion of mediated effect was calculated by the formula of (beta1*beta2)/beta0. P-value of <0.05 was considered as statistical significance. To account for multiple testing correction, Bonferroni correction was also used to adjust the thresholds of significance level, with a strong evidence of P<0.05/(number of genes) and a suggestive evidence of 0.05/(number of genes)<=P<0.05. All analyses were conducted by using SMR software (version 1.03) or R software (version 4.1.0).

## Results

### Genetic instrument selection

As shown in **Figure 1** **and Supplementary Table 2,** a total of 7,737 SNPs significantly associated with the expression in blood of 17 genes encoding PDE1 to PDE9 were identified. While there is a lack of available cis-eQTLs identified for PDE10 and PDE11, thus these two PDEs were not included in the analyses. The most significant cis-eQTL for all 17 encoded genes had F statistics of >10, indicating a small role of weak instrument bias in our study. Approved or investigational drugs targeting PDEs are presented in **Supplementary Table 2.**

### MR analysis between the expression of genes encoding PDEs in blood and outcomes

As shown in **Figure 2** **and Supplementary table 3**, the most significant association with the risk of both CHD and MI was found on PDE5A expression, in which a 1-SD increase of PDE5A expression was associated with increased risk of CHD with an OR of 1.22 (95% CI=1.06-1.40, PLJ=LJ0.0046) or MI with an OR of 1.27 (95% CI=1.09-1.48, PLJ=LJ0.0024). The observed association with MI kept significant after bonferroni correction using a significance threshold of PLJ<LJ0.003 (multiple testing for 17 genes) and HEIDI test suggested a small role of linkage disequilibrium in the observed association (P_CHD_=0.160; P_MI_=0.375). Besides, MR analysis provided a suggestive evidence for a positive association between PDE8A expression and the risk of CHD (OR=1.26, 95% CI=1.07-1.49, PLJ=LJ0.0047) and MI (OR=1.24, 95% CI=1.04-1.48, PLJ=LJ0.0175), which tended to null after bonferroni correction and HEIDI test in MI analysis showed the potential presence of linkage disequilibrium (P<0.01). As shown in **Supplementary Table 5**, six genes were identified nearby the instrumental variant of PDE5A expression in blood (rs10518336) and the expression of which were significantly related to the instrumental variant. Among them, the expression of four genes were associated with the risk of outcomes, indicating that the horizontal pleiotropy may not be ruled out **(Supplementary Table 6)**. No association was observed between the expression of gene nearby the instrumental variant of PDE8A expression in blood (rs67804993) and the outcomes **(Supplementary Table 6)**.

**Figure 2.**
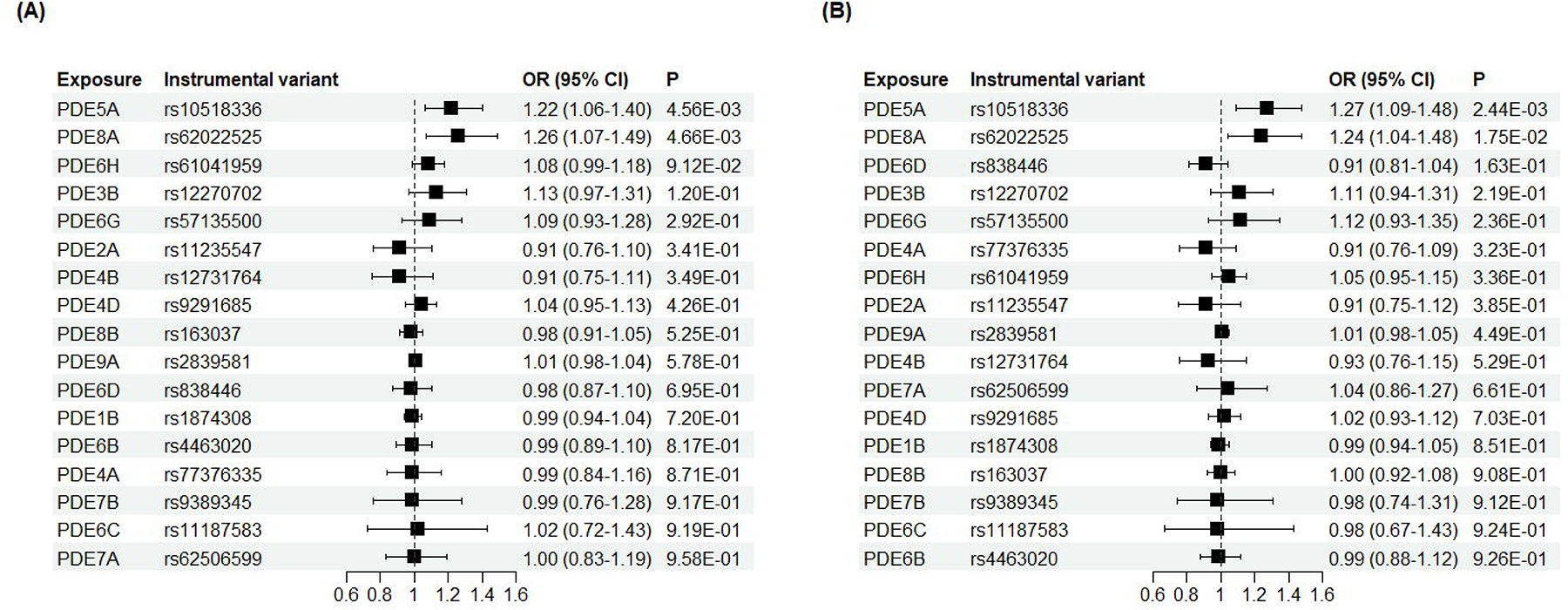
Association between the expression of PDEs in blood and coronary heart disease or myocardial infarction. (A) Coronary heart disease; (B) Myocardial infarction. * Shown in ascending order of p value.

### MR analysis between the expression of PDE5A or PDE8A in artery or heart tissues and outcomes

As MR analyses found a strong evidence for the association between the expression of PDE5A or PDE8A and the risk of CHD and MI, we further examine the association of PDE5A/PDE8A expression in relevant tissues with outcomes. Heart and artery are the predominating organs in cardiovascular diseases. **Supplementary** Figure 1 and 2 display the tissue-specific expression of PDE5A and PDE8A from GTEx Portal. PDE5A was expressed highest in artery aorta, followed by artery tibial, and the expression in artery coronary ranked six place, indicating an important role of PDE5 in artery. While PDE8A was expressed highest in nerve tibial, and the expression in artery tibial ranked 15 place.

There were 188, 13 and 9 cis-eQTLs identified to generate instruments for PDE5A expression in artery aorta, artery tibial and heart atrial appendage respectively, and 2 cis-eQTLs were identified as instruments for PDE8A expression in artery tibial (**Figure 1**). As shown in **Figure 3** **and Supplementary table 4**, the associations between PDE5A expression and the outcome were also found in artery aorta (OR_CHD_=1.17, 95% CI=1.05-1.32, PLJ=LJ0.0066; OR_MI_=1.15, 95% CI=1.02-1.30, PLJ=LJ0.0269) and heart atrial appendage (OR_CHD_=1.20, 95% CI=1.02-1.41, PLJ=LJ0.0312; OR_MI_=1.18, 95% CI=0.99-1.40, PLJ=LJ0.0667). The observed association with CHD passed the bonferroni correction test (PLJ<LJ0.013, multiple testing for 4 genes) and the HEIDI test (P>0.01). No nearby genes were found to be significantly related to the instrumental variant of PDE5A expression in artery aorta (rs1393966) (**Supplementary table 5**). No association was observed between the expression in heart atrial appendage of gene nearby the instrumental variant of PDE5A expression (rs75098287) and the outcomes **(Supplementary Table 6)**. These results suggesting a small role of the horizontal pleiotropy in the observed associations.

**Figure 3.**
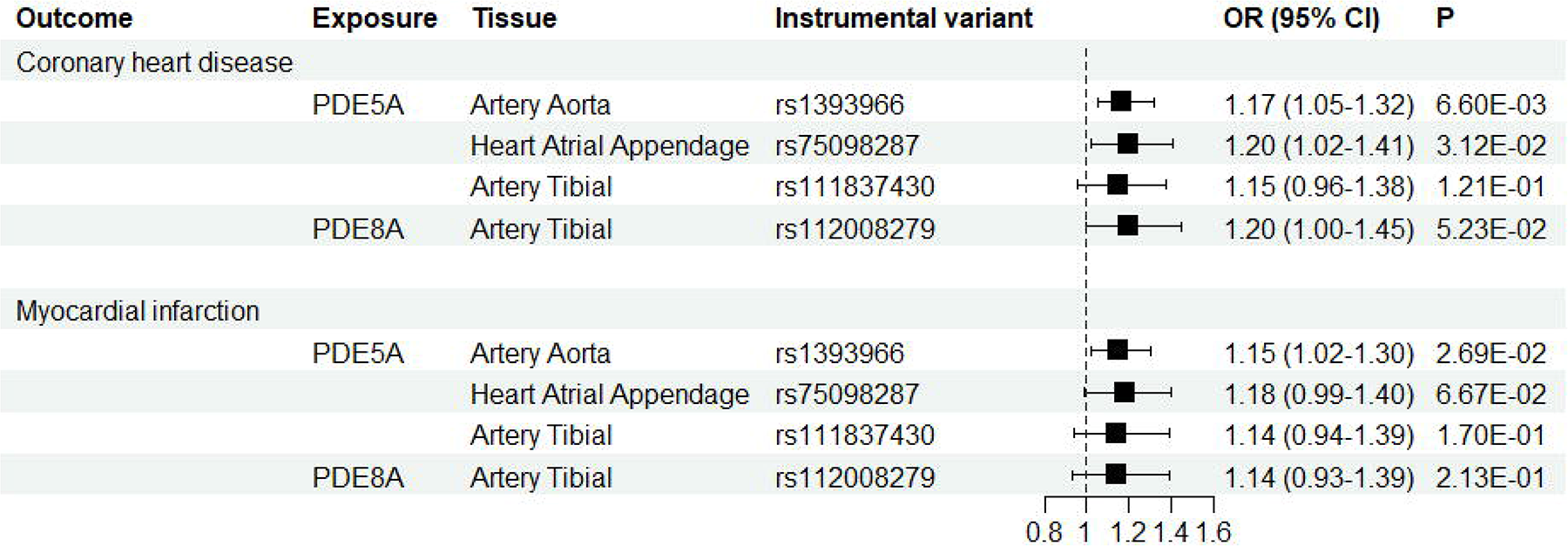
Association between the expression of PDE5A or PDE8A in artery or heart tissue and coronary heart disease or myocardial infarction.

### MR analysis between the methylation of PDE5A or PDE8A in blood and outcomes

There were 1,185 and 310 cis-mQTLs in blood identified to generate instruments of DNA methylation in 14 CpG sites of PDE5A and 12 CpG sites of PDE8A, respectively (**Figure 1**). Among these CpG sites, the methylation level of 12 CpG sites of PDE5A and 7 CpG sites of PDE8A robustly associated with the expression of the corresponding gene had significant associations with the risk of CHD and MI in the same direction, indicating that methylation in these CpG sites may affect the outcomes via regulating gene expression(**Figure 4, Supplementary table 7 and 8**).

**Figure 4.**
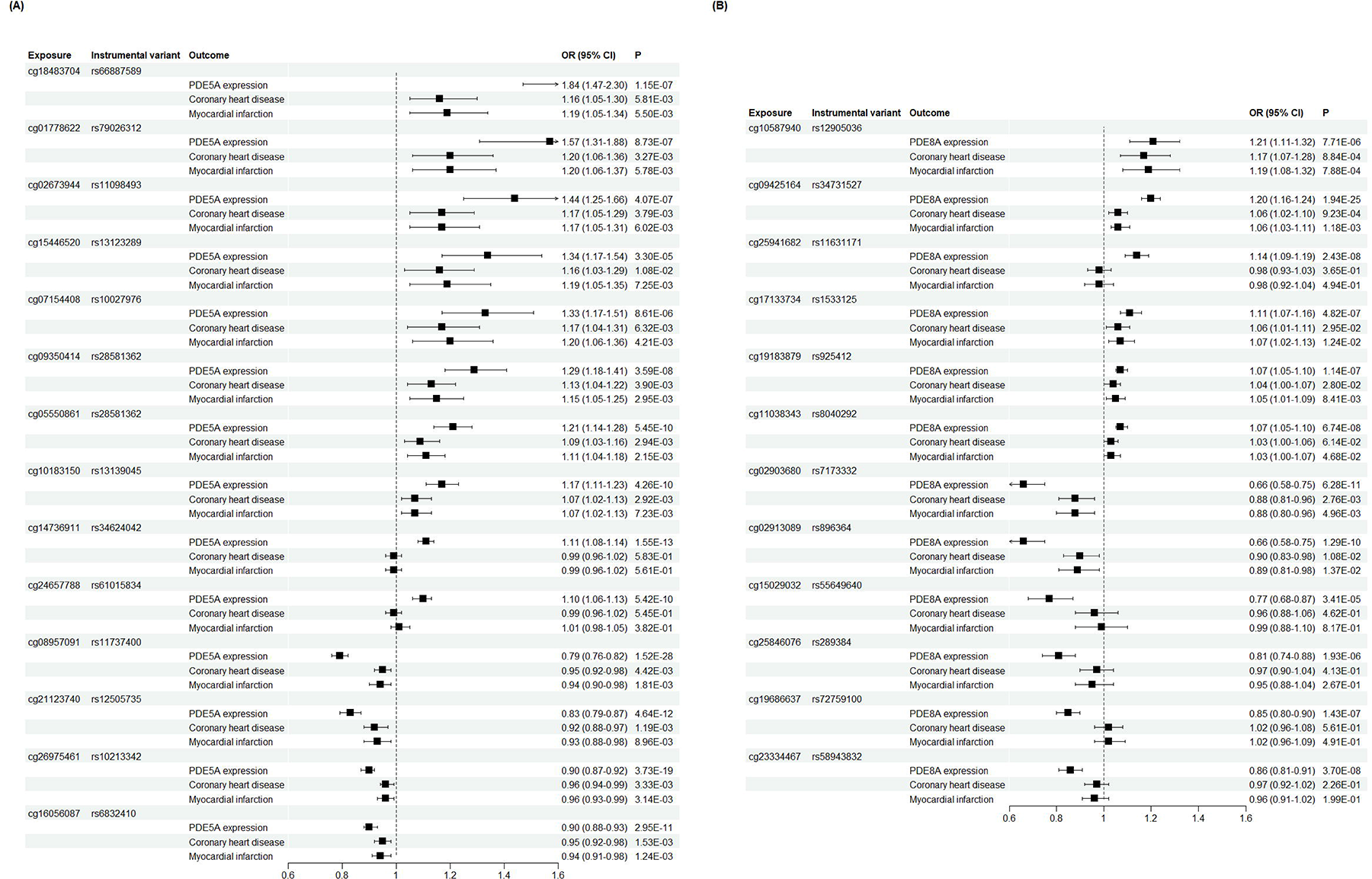
Association between the methylation of PDE5A or PDE8A and the gene expression in blood, coronary heart disease or myocardial infarction. (A) PDE5A; (B) PDE8A.

### Two step MR analyses between PDE5A expression, metabolic traits and outcomes

As shown in **Supplementary table 9,** the expression of PDE5A but not PDE8A was related to multiple metabolic traits, thus two step MR analyses were performed to explore the causal relationship between PDE5A expression, metabolic traits and the risk of CHD or MI. **Figure 5** showed that the association between PDE5A expression and CHD or MI was partly mediated by diastolic blood pressure (Proportion-mediated _CHD_=25.97%; Proportion-mediated _MI_=21.42%), systolic blood pressure (Proportion-mediated _CHD_ =15.77%; Proportion-mediated _MI_ =11.63%) and LDL-C (Proportion-mediated _CHD_=15.35%; Proportion-mediated _MI_ = 8.17%). The association between PDE5A expression and MI was mostly mediated by CHD (Proportion mediated=78.84%). No mediated role of body mass index, blood glucose or other lipids was observed between PDE5A expression and outcomes.

**Figure 5.**
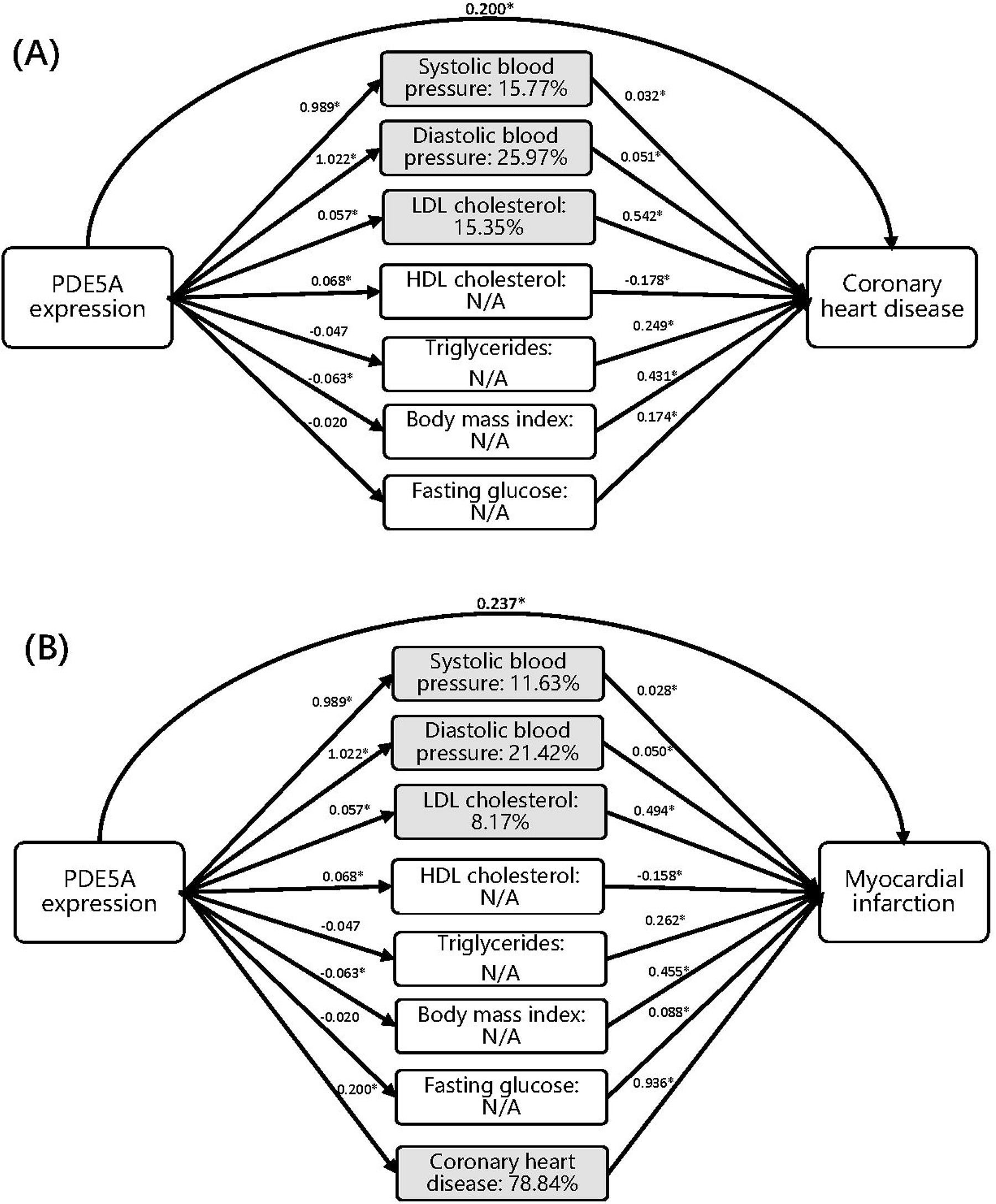
Two-step mendelian randomization between PDE5A expression, metabolic traits and coronary heart disease or myocardial infarction. (A) Coronary heart disease; (B) Myocardial infarction. * Percentage in plots indicates the proportion of mediated effect. The asterisk in plots indicates statistical significance (P<0.05).

## Discussion

To investigate the therapeutical potential of PDE inhibitors for ischemic heart disease and the underlying pathways, we explored the association of the expression of genes encoding PDEs (i.e., PDE1-PDE9) with the risk of CHD and MI in this MR study. Two types of PDEs, PDE5 and PDE8, were identified to have the potential to be repurposed for CHD or MI prevention in this study. Specifically, a strong evidence was observed for the positive association of PDE5A expression in blood and artery aorta (the tissue with highest expression) with the risk of CHD and MI. We also identified 12 CpG sites, the methylation level of which was significantly related to the risk of CHD and MI via affecting the PDE5A expression. Mediation analysis suggested that PDE5 inhibition might reduce the incidence of ischemic heart disease partly through regulating blood pressure and LDL cholesterol. These findings provided genetic evidence that PDE5 inhibitors may be effective in the prevention of CHD and MI, especially in protecting against MI among patients with CHD.

PDE5, encoded by PDE5A gene, is an enzyme selectively hydrolyzing cGMP, which is distributed across most tissues of human body. Thus, it is known that PDE5 inhibitors, including sildenafil, vardenafil and tadalafil, exert its therapeutic effect by inhibiting cGMP degradation. PDE5 inhibitors were originally developed to treat angina while failed in clinical trials, then repurposed and approved by FDA for treating erectile dysfunction in 1998, pulmonary arterial hypertension in 2005 and benign prostatic hyperplasia in 2011 (4–6). Given of the safety of PDE5 inhibitors and the involvement of cGMP in vast physiological processes, enormous interest has been raised to explore the potential new functions of PDE5 inhibitors(6).

Growing preclinical studies have reported that PDE5 inhibitors play a protective role against cardiovascular diseases, such as heart failure, MI, ischemia-induced ventricular arrhythmias, myocardial hypertrophy and so on (4–6). Several clinical trials have demonstrated the benefits of PDE5 inhibitors in patients with heart failure, while the effect on ischemic heart disease is still uncertain (28). A number of observational studies have reported the protective effect of PDE5 inhibitors on ischemic heart disease (29–31). For example, a nationwide Danish cohort study including 71,710 men found that PDE5 inhibitors use was related to a decreased incidence of MI as compared to the general male population without receiving PDE5 inhibitors (RR=0.74; 95% CI=0.69-0.79), however indication bias may play a role in the observed association (31). Another case-control study including type 2 diabetic men with silent coronary heart disease found that treatment with PDE5 inhibitors and statins was negatively associated with the occurrence of major adverse cardiac events (HR=0.66, 95%CI=0.46-0.97), while in which the effect of statins and PDE5 inhibitors cannot be distinguished and indication bias cannot be eliminated as well(30). A recent real-world study based on Swedish nationwide registers investigated the protective effect by comparing the survival between men treated with PDE5 inhibitors and those treated with alprostadil in order to reduce the influence from indication bias, since erectile dysfunction is the major indication of PDE5 inhibitors and alprostadil is another treatment for erectile dysfunction(29). This study included 18,542 men with stable coronary heart disease and found that men treated with PDE5 inhibitors had a lower mortality as well as a reduced incidence of major adverse cardiac events, including MI (HR= 0.81, 95%CI=0.70–0.93). Although indication bias can be reduced as much as possible by using active comparator, it is impossible to be completely avoided and unknown confounders cannot be ruled out in observational studies. Besides, as these drugs were mainly prescribed for men to treat erectile dysfunction in real world, it is unlikely to assess the effect in general population. Our study observed consistent results that increased expression of PDE5A was related to a higher risk of CHD and MI, which provided strong supportive evidence for findings from previous observational studies. Besides, we found that the mediated proportion of CHD on the positive relation between PDE5 and MI risk was as high as round 80%, indicating that PDE5 inhibition probably contributed to preventing patients with CHD from MI. Furthermore, PDE5 has been emerged as novel target for treating metabolic disorders, which are known risk factors of ischemic heart disease (18). This study also observed a mediated effect of blood pressure and LDL cholesterol between PDE5 inhibition and ischemic heart disease, which offered some clues for the underlying mechanisms, calling for further studies.

PDE8 was also identified in this study, the inhibition of which was recently found to be a potential novel therapeutic target of treating airway diseases and dementia (32,33). No studies reported the cardiovascular effect of PDE8. Although this MR study found that the expression in blood of PDE8A was related to the risk of CHD and MI, the association did not pass the bonferroni correction test. Besides, PDE8A was expressed less in cardiovascular organs and no relationship was observed between the PDE8A expression in artery and the outcomes. Therefore, whether PDE8 inhibition really had a cardiovascular benefit need to be further investigated.

This is the first MR study to comprehensively explore the causal effects of PDEs on ischemic heart disease. The use of QTLs as instruments to proxy drug exposure in MR study could overcome the shortcomings of observational studies, such as confounding bias and reverse causation. Besides, several types of QTLs (eQTLs in blood, artery aorta and heart, mQTLs in blood) have provided consistent evidence, indicating the robustness of the protective effect of PDE5 inhibition on ischemic heart disease. Two-step MR analysis contributed to identifying the subgroup population that may benefit more from PDE5 inhibitors use. However, there are several limitations. Firstly, this study used the expression of drug target genes to proxy drug exposure, which might not be equivalent to clinical treatment because of a gap between drug targets and available agents, for example, there may be various effects across different types of PDE5 inhibitors. However, these findings could provide additional evidence for previous observational studies regarding the preventive effect of PDE5 inhibitors against ischemic heart disease. Hence, RCTs are required to further investigate the efficacy of different PDE5 inhibitors. Secondly, horizontal pleiotropy cannot be excluded when using eQTLs in blood as instruments for PDE5A expression in primary analysis, while consistent associations were observed using eQTLs in tissues or mQTLs in blood as instrument, suggesting the robustness of the findings. Thirdly, false negative association of other PDEs cannot be ruled out in drug target MR study as there may be unidentified eQTLs which play a more important role in gene expression than the identified top eQTL. Fourthly, this study was conducted based on summary-level data, thus we were unable to explore the specific effect in subgroup population, such as in men and women. Fifthly, genetic data of this study were mainly collected from population of European ancestry, leading to a limited generalization of these findings.

## Conclusion

In conclusion, this drug-target MR study identified PDE5 that had the potential to be repurposed as novel target for CHD and MI prevention, especially in protecting against MI among patients with CHD, and the underlying mechanisms of the effect might be partly involved in regulating blood pressure and LDL cholesterol.

## Data Availability

All raw data produced are available online at eQTLGen Consortium, GTEx Portal, Global Lipids Genetics Consortium, International Consortium of Blood Pressure, GIANT Consortium, MAGIC Consortium and CARDIoGRAMplusC4D Consortium.

## Abbreviations

CHD: coronary heart disease
CI: confidence interval
CpG: Cytosine-guanine dinucleotides
eQTLs: expression quantitative trait loci
GWAS: genome-wide association study
HEIDI: heterogeneity in dependent instruments
mQTL: Methylation quantitative trait loci
MI: myocardial infarction
OR: odds ratio
PDE: phosphodiesterase
SNP: single-nucleotide polymorphism
SMR: summary-data-based Mendelian Randomization

## Acknowledgments

The data used for the analyses described in this manuscript were obtained from the eQTLGen Consortium, GTEx Portal, Global Lipids Genetics Consortium, International Consortium of Blood Pressure, GIANT Consortium, MAGIC Consortium and CARDIoGRAMplusC4D Consortium on 01/07/2023. We thank the patients and investigators who contributed to these studies.

## Ethical statement

All publicly available genome-wide association studies (GWASs) and quantitative trait loci (QTLs) studies used in this MR analyses were approved by the relevant institutional review boards with informed consents from participants.

## Funding Information

This work was supported by National Natural Science Foundation of China (82304213) and Start-up Fund for high-level talents of Fujian Medical University (XRCZX2021026) to Dr. Wuqing Huang. The funder had no role in study design, data collection and interpretation, or the decision to submit the work for publication.

## Author Contributors

All authors were responsible for the study concept and design. WH obtained funding. JX did the statistical analysis. JX and WH drafted the manuscript, and all authors revised it for important intellectual content.

## Competing interests

The authors declare no conflict of interest.

## Notes

### Competing Interest Statement

The authors have declared no competing interest.

### Funding Statement

This study was funded by National Natural Science Foundation of China (82304213) and Start-up Fund for high-level talents of Fujian Medical University (XRCZX2021026) to Dr. Wuqing Huang. The funder had no role in study design, data collection and interpretation, or the decision to submit the work for publication.

### Author Declarations

The study used ONLY openly available human data that were originally located at eQTLGen Consortium, GTEx Portal, Global Lipids Genetics Consortium, International Consortium of Blood Pressure, GIANT Consortium, MAGIC Consortium and CARDIoGRAMplusC4D Consortium.

