## Supplementary figures and images for "Combination of QTL and GWAS to uncover the role of phosphodiesterases in ischemic heart disease"

### Supplementary figure 1

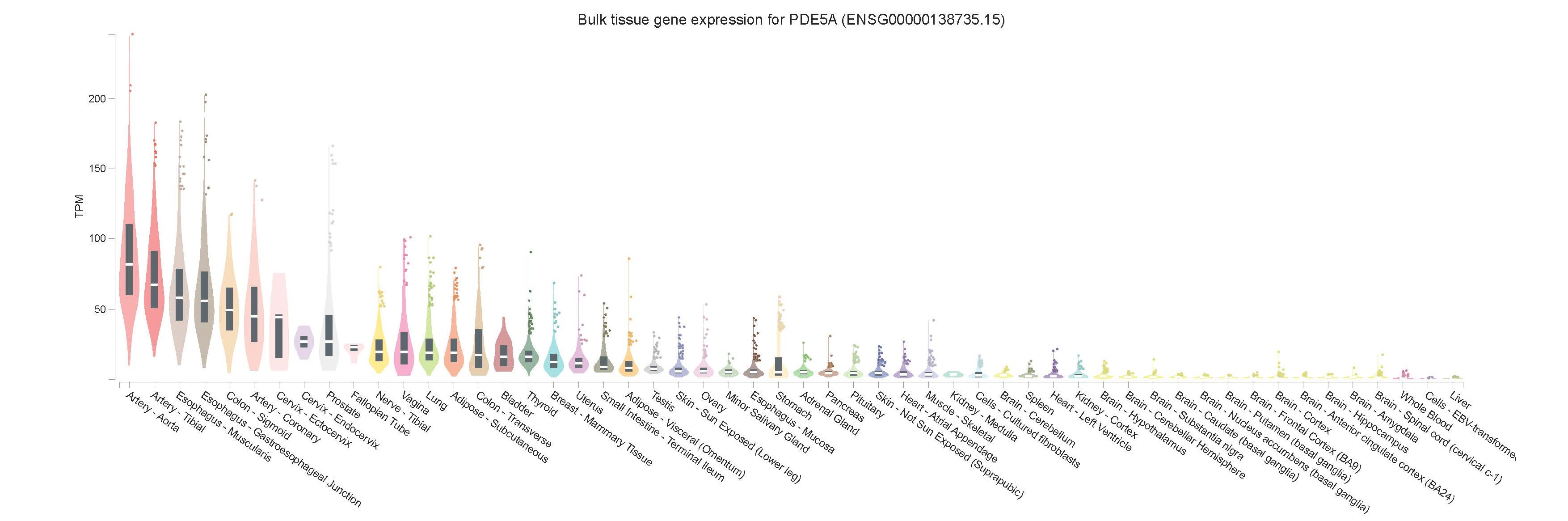

### Supplementary figure 2

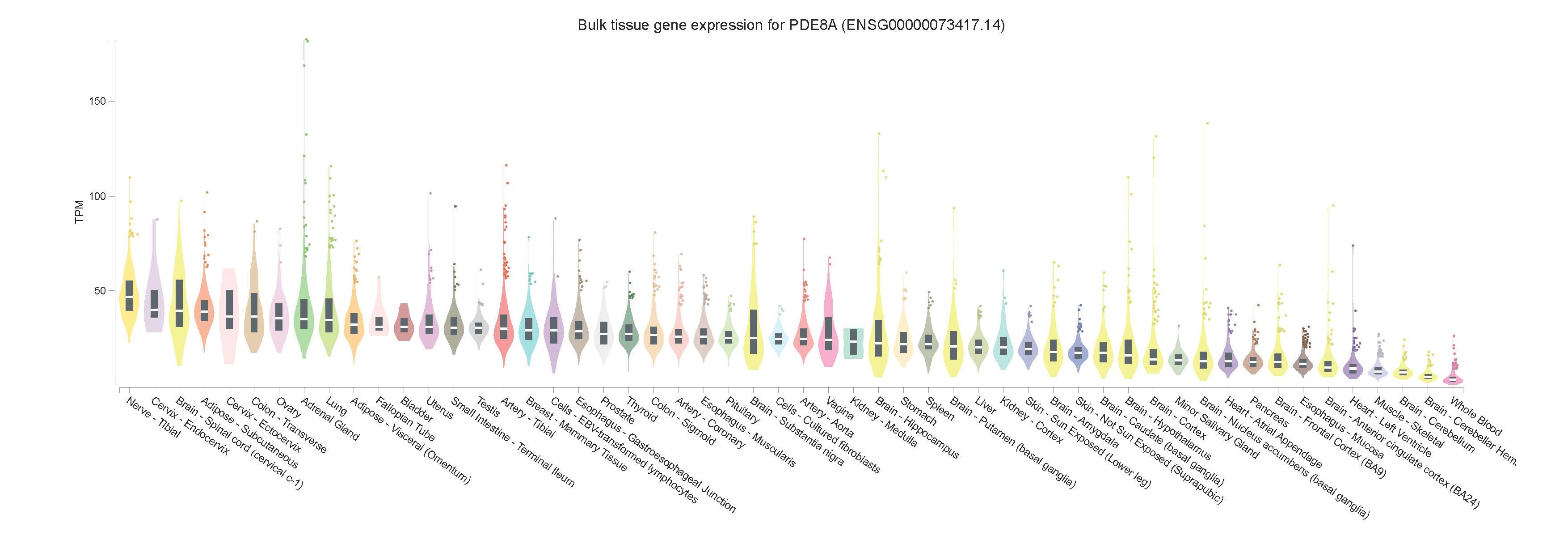
